# Awareness and perceptions of social prescribing among university students in the UK

**DOI:** 10.64898/2026.07.07.26357397

**Authors:** Jessica K Bone, Daisy Fancourt, Daniel Hayes

**Affiliations:** Research Department of Behavioural Science and Health, Institute of Epidemiology & Health Care, University College London, UK

**Keywords:** Social prescribing, University, Students, Extracurricular, Community

## Abstract

Universities provide a key opportunity to deliver social prescribing, a care pathway that aims to connect people with non-medical forms of support within the community to address their social, emotional, and practical needs. However, it is unclear whether students in the UK are aware of social prescribing and whether it would be an acceptable form of support. We surveyed 775 university students across the UK who completed a questionnaire measuring awareness and perceptions of social prescribing. We described awareness and attitudes and used logistic regression to explore how they differed according to individual characteristics. We found an awareness-attitude paradox. Only 25% of students were aware of social prescribing, but attitudes were overwhelmingly positive once explained: 97% thought it could support mental health and wellbeing; 95% believed universities should offer it; and 89% would accept social prescribing if offered by a healthcare professional. Students who were older, postgraduates, and had English as their first language were among those with higher odds of being aware of social prescribing, but positive attitudes were more evenly reported across the sample. Our findings indicate that implementation efforts should prioritise awareness-raising and clear referral pathways, rather than increasing students’ willingness to engage with social prescribing.

## Introduction

The prevalence of psychological distress and mental health disorders has markedly increased among students in higher education in recent years.^1,2^ Between 25% and 35% report symptoms indicative of a mental disorder.^3^ This can lead to poorer academic performance and increased risk of dropping out of university.^2,4^ For many students, university is a new setting that brings a novel set of financial, academic, and social stressors.^5,6^ This context can also make students less likely to exercise and do other leisure activities, restricting their focus to the academic work that is often the catalyst for stress.^5,7–10^ On top of this, students also report difficulties in trying to navigate fragmented and complex health services whilst at university,^11^ leading many to try and self-manage health problems.^12^ Despite these known psychological challenges, only 17% of students with a likely mental disorder have used mental health services in the previous 12 months.^12^ Furthermore, those who do seek support are often subject to long waiting lists.^11^ Collectively, this situation highlights the need to consider other ways of supporting the mental health of students.

In the United Kingdom (UK), there is increasing focus on using community-based approaches to prevent poor health.^13^ Social prescribing (SP), a care pathway that aims to connect people with non-medical forms of support within their community,^14^ is central to this. SP targets people’s social, emotional, and practical needs, identified through conversations about what matters to them.^15^ In the UK, SP typically consists of referral to a Link Worker, or other similar professional, who works with the individual to develop a personalised care plan that connects them to community support. This may include the arts, sports, nature-based activities, volunteering, counselling, mentoring, skills and development opportunities as well as broader advice and information, such as debt advice and housing support.^16^ Emerging evidence indicates that SP may reduce anxiety and depression, enhance wellbeing, and reduce other healthcare utilisation.^17–22^

Universities provide a key opportunity to deliver SP. They offer the ideal setting for a whole system approach to support health and wellbeing, as set out in the Healthy Universities framework^23^ and the University Mental Health Charter.^24,25^ A wide range of academic, social, and leisure activities, health services, counselling, financial support, and student-led groups are usually on offer within universities, as well as in the wider community, providing ample opportunities for students to be referred to. SP thus presents an accessible, scalable, and non-stigmatising care pathway that could help connect students to existing resources. However, despite emerging evidence for the benefits of SP and associated activities, including amongst adolescents and young adults, it has not yet been implemented and evaluated in university settings.^25,26^

A recent scoping review found that the existing evidence on SP in university settings is concentrated almost exclusively within two areas, namely educating the future healthcare workforce about SP and involving students in delivering SP services, with little research examining students as recipients of SP.^26^ NHS England and the National Academy for Social Prescribing (NASP) established the SP Student Champion Scheme in 2016, aiming to embed SP within future health professionals’ education.^27,28^ During this initiative, awareness of SP increased from 7% to 75% of sampled medical students in universities with champions.^29,30^ Additionally, once educated about SP, over 95% felt SP was a useful concept that would be relevant to their future practice and should be part of the undergraduate medical curriculum.^29,30^ However, to date, only one study has explored attitudes towards SP in the general student population.^31^ Findings were similar to those from UK medical students but this study focused on Canadian university students. It thus remains unclear whether university students in the UK are aware of SP and whether it would be an acceptable and useful addition to services.

Understanding students’ attitudes towards SP is vital for its development in universities. The University Mental Health Charter highlights that student mental health services must be coproduced, using the student voice to ensure that services can meet the changing needs of the diverse student body.^24,32^ Additionally, awareness and acceptability are important precursors to successful implementation of any new health interventions.^33^ Low awareness may limit engagement, uptake, and equitable reach, even where an intervention is viewed positively once explained. Understanding both awareness and acceptability is therefore an important first step in assessing implementation readiness and identifying whether future implementation efforts should prioritise increasing awareness, improving intervention design, or addressing attitudinal barriers. Consequently, in this study, we aimed to 1) explore awareness of and attitudes towards SP among UK university students; and 2) describe how awareness and attitudes differed across sociodemographic groups.

## Methods

### Sample

The target population was UK university students. Adverts were shared via social media and the authors’ national networks, including the Social Prescribing Youth Network (SPYN) and academics in university settings. The survey was completed between June 2024 and January 2025. A total of 1060 participants individuals accessed the survey, 865 (82%) of whom gave informed consent. To be eligible, individuals then had to confirm that they were an undergraduate or postgraduate (including doctoral) student in the UK. Of those who consented, 859 (99%) were students, 777 (90%) of whom reported that they attended a UK university. We excluded participants who had two or more responses that indicated suspicious bot-like free text responses, such as random letter sequences and Latin placeholder phrases (n=2, <1%). This left a final analytical sample of 775 UK university students.

### Ethical approval

This study was approved by University College London Research Ethics Committee (project 6735/018). All participants gave informed consent, and all research was performed in accordance with the Declaration of Helsinki.

### Measures

#### Demographics

Participants self-reported their age (years), gender (male [including trans man], female [including trans woman], non-binary/other), ethnicity (White, Asian/Asian British, Black/African/Caribbean/Black British, mixed/multiple ethnic groups, other), and first language (English, not English). They also reported on their student status, including whether they were an international student, level of study (undergraduate, masters, doctoral), year of study, and academic field. Academic field included five categories: humanities (languages, historical philosophical and religious studies, design, creative and performing arts); social science (anthropology, economics, geography, psychology, sociology); natural sciences (biology, chemistry, physics, earth science, astronomy); maths/computing (formal science); and applied sciences/professions (medicine and allied subjects, engineering, architecture, law, business, etc). Year of study was used for descriptive statistics only due to issues with perfect prediction.

#### Social determinants of health

The Social Needs Screening Tool measured five core health-related social needs, including housing, food, transportation, utilities, and personal safety, as well as the additional needs of employment, childcare, and financial strain.^34^ We calculated a total score as a sum of the number of social needs, with higher scores indicating greater needs (total possible range 0-9).

#### Health and wellbeing

We used binary indicators of whether participants had any 1) physical disabilities or long-term physical health problems and 2) mental health difficulties or diagnosable mental health conditions. The short Warwick-Edinburgh Mental Wellbeing Scale (SWEMWBS) measured mental wellbeing, with raw scores transformed to metric scores on the interval scale, and higher scores indicating greater mental wellbeing (total possible range 7-35).^35^

#### Health behaviours

Participants reported for the last three months: how often they had done things to look after their mental health (never, rarely, sometimes, often, always); whether they had spoken to a health professional about their mental health and wellbeing (yes, no); whether they had used other sources of support for their mental health and wellbeing (none, family/friend, other source); and whether they had taken pills or medication to help with their mental health and wellbeing (yes, no). Finally, they reported whether in the last three months, because of their mental health and wellbeing, they had missed any of their university course (yes, no) or employed work (yes, no).

#### Social prescribing

Measures of SP awareness and attitudes were designed following previous research.^29,31^ Before giving participants any information about what SP involves or how it is defined, we measured awareness using one item: “Have you heard about social prescribing before?”, to which participants could respond yes, no, or I am not sure. If they responded yes, participants used free text to report where they heard about SP. We grouped the free text responses into five categories based on frequency: participants heard about SP through 1) teaching, 2) healthcare, 3) family/friend, 4) work, and 5) other. Other included in the news, at events, through research, and on social media, among other places.

Participants were then presented with a detailed definition and example of SP (see Supplementary Materials), based on the internationally accepted definition,^14^ after which we measured prior engagement in SP. Participants reported whether they had taken part in any social or community activities (e.g. arts group, music/singing group, book/writing group, volunteering, social club, sports club, nature activity, etc.) with the aim of improving their health and wellbeing in the last 3 months (yes, no). Those who had done so then indicated whether these activities were accessed through a social prescriber/link worker/community connector/care navigator (yes, no).

Additionally, a series of five questions assessed students’ positive attitudes towards SP. These measured whether participants 1) think SP could support mental health and wellbeing, 2) think their university should offer SP, 3) would be willing to follow a healthcare professional’s advice if they were to suggest SP at an appointment, 4) think that SP is relevant to future health professionals (e.g. nursing students, medical students) as an intervention they can offer patients, and 5) think that SP should be part of the curriculum for health sciences programs (e.g. nursing school, medical school, public health, medical sciences). All questions had binary response options (yes, no). As there was little variation in attitudes towards SP, we created a binary indicator of whether participants were entirely open to and positive towards SP (agreeing with all 5 items) or had fewer positive attitudes towards SP (agreeing with 0-4 items).

### Statistical analysis

We first explored awareness of and attitudes toward SP descriptively. Following this, we tested which individual characteristics (socio-demographics, health, health behaviours) were associated with awareness of and attitudes towards SP in logistic regression models. For both outcomes, we applied a series of regression models containing similar predictors in blocks, accounting for confounders relevant to each block to avoid potential “table 2 fallacy”.^36^ Model 1 included all sociodemographic factors. Model 2 additionally tested health-related factors, adjusting for sociodemographic factors as they likely confound the association between health-related factors and SP awareness/attitudes. Model 3 then tested associations with health behaviours, adjusted for all variables from Model 2. For attitudes towards SP, a final model 4 additionally included awareness of SP.

For participants with missing responses, we imputed data using multiple imputation by chained equations (MICE).^37^ We used truncated linear, logistic, ordered logistic, and multinomial logistic regression according to variable type, generating 20 imputed datasets (maximum missing data 6%; Table S1). The imputation models included all variables used in analyses, but no auxiliary variables were available. Findings from imputed analyses did not differ to complete case analyses (Tables S6-S8), so results are reported following imputation.

Our main analyses were unweighted. For comparison in supplementary analyses, we generated probability weights to make the sample representative of university students in the UK. We weighted the final analytical sample to match the Higher Education Statistics Agency (HESA) official student statistics for 2022/2023^38^ according to gender, age, ethnicity, and level of study, using the Stata *ebalance* package.^39^ To do this, we had to exclude participants missing data on weighting variables (<1%; Table S1) as well as participants of non-binary/other gender as they were not represented in the HESA statistics (2%). To remove extreme variation, weights were capped at the 95^th^ percentile and normalized so that the total summed to the number of participants in the weighted sample (n=761). All analyses were performed using Stata 18.^40^

## Results

We included 775 students aged between 18 and 75. Before weighting, the mean age was 24.53 (standard deviation [SD]=7.66, median=22) with 76% female, 22% male, and 2% non-binary or other gender (Table 1). Overall, 67% were White and 88% had English as a first language. Most were undergraduates (77%) in their first year (37%) and were not international students (83%). On average, students reported a total of 2.59 social needs (SD=2.13), with difficulties most frequently reported in the housing, food, and financial strain domains.

**Table 1.** Sample characteristics. *Note.* N=775 before weighting, N=761 after weighting; participants missing ethnicity data and those of non-binary/other gender excluded when generating population weights.

|  | Unweighted | Weighted |
| --- | --- | --- |
|  | Mean (SD) |  |
| Age | 24.53 (7.66) | 25.58 (9.29) |
| Social needs (range 0-9) | 2.59 (2.13) | 2.66 (2.10) |
| Mental wellbeing (SWEMWBS, range 7-35) | 20.33 (4.14) | 20.39 (4.08) |
|  | Proportion |  |
| Gender |  |  |
| Male | 22% | 41% |
| Female | 76% | 59% |
| Non-binary/other | 2% | - |
| Ethnicity |  |  |
| White | 67% | 71% |
| Asian/Asian British | 16% | 14% |
| Black/African/Caribbean/Black British | 5% | 8% |
| Mixed/multiple ethnic groups | 10% | 5% |
| Other | 2% | 2% |
| First language English | 88% | 89% |
| International student | 17% | 16% |
| Level of study |  |  |
| Undergraduate | 77% | 71% |
| Masters | 19% | 23% |
| Doctoral | 4% | 6% |
| Year of study |  |  |
| 1 | 37% | 37% |
| 2 | 25% | 26% |
| 3 | 22% | 21% |
| 4 | 13% | 12% |
| 5 | 2% | 2% |
| Other | 1% | 2% |
| Academic field |  |  |
| Social science | 37% | 38% |
| Humanities | 11% | 12% |
| Natural sciences | 10% | 12% |
| Maths/computing (formal science) | 9% | 9% |
| Professions and applied sciences | 32% | 29% |
| Physical health problems or disabilities | 17% | 15% |
| Mental health difficulties or conditions | 34% | 31% |
| Done things to look after mental health |  |  |
| Never | 3% | 5% |
| Rarely | 16% | 17% |
| Sometimes | 40% | 40% |
| Often | 31% | 29% |
| Always | 10% | 10% |
| Spoken to professional about mental health | 42% | 41% |
| Other mental health support |  |  |
| None | 33% | 34% |
| Family/friend | 64% | 63% |
| Other | 3% | 3% |
| Taken medication for mental health | 22% | 21% |
| Missed university due to mental health | 30% | 28% |
| Missed employed work due to mental health | 10% | 8% |

A large proportion of participants were from the University of Plymouth (60%) and University College London (14%). Despite this, our sample broadly aligned with the UK university student population, 71% of whom are White and 70% undergraduates. However, females were over-represented (76% of our sample vs 57% nationally) as were those studying social sciences (37% vs 8% nationally) and professions/applied sciences (32% vs 15% nationally), leaving those studying humanities under-represented (11% vs 58% nationally).^38^ After weighting, sample characteristics were similar to the UK university population in terms of gender, age, ethnicity, and level of study (Table 1).^38^

### Awareness of social prescribing

Overall, only 25% of students had previously heard of social prescribing, with another 8% unsure whether they had heard of it (Figure 1). For those who were aware of SP, most were familiar with it through teaching (39%), other sources (24%), or work (16%). Fewer had heard about SP from healthcare (13%) or family/friends (8%). Overall, 36% had taken part in social or community activities to improve their health and wellbeing in the last 3 months, but only 14% of these students did so through a social prescriber/link worker/community connector/care navigator (n=37).

**Figure 1.**
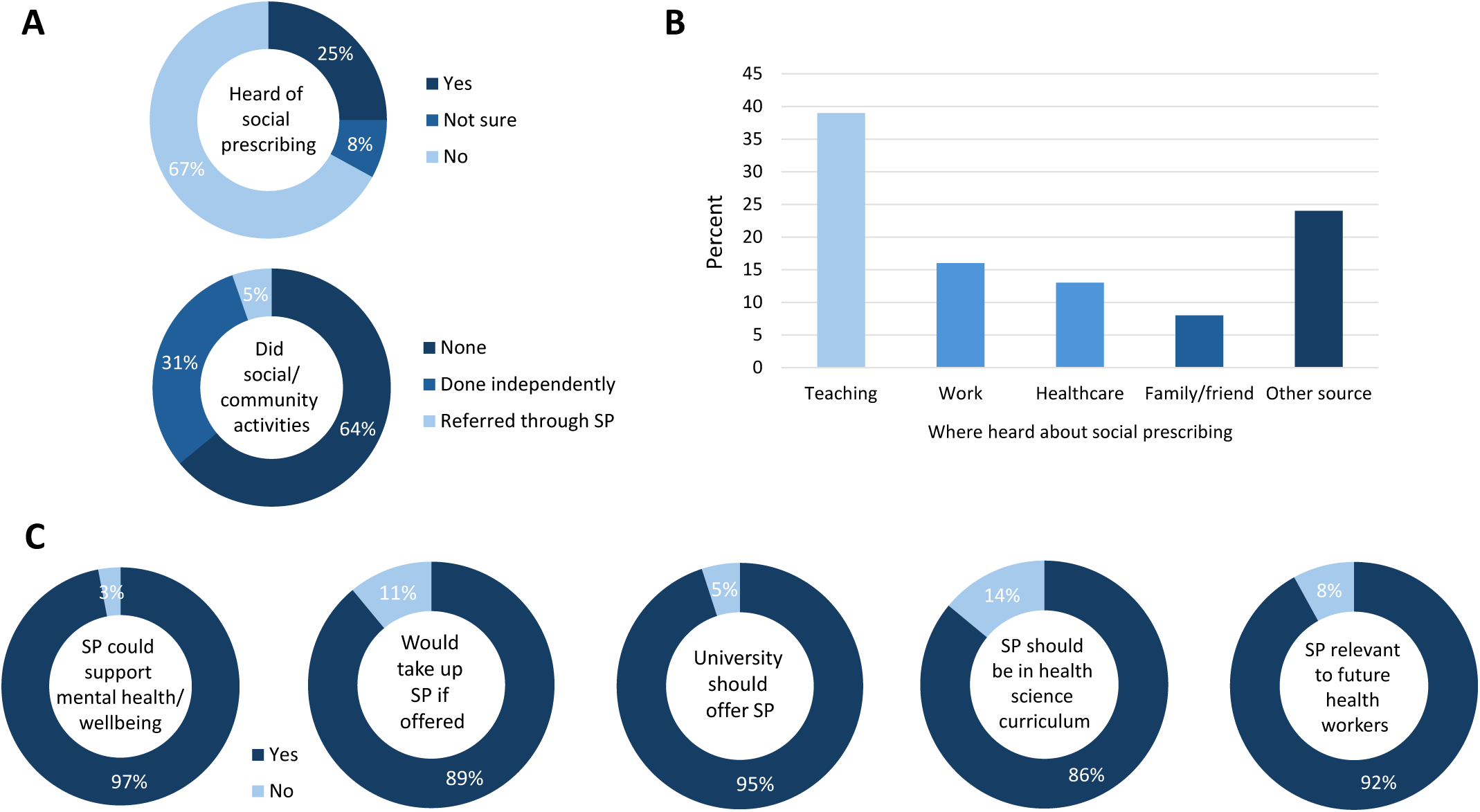
Descriptive statistics for social prescribing measures. A: Awareness and use of social prescribing (n=775). B: Source where students previously heard about social prescribing (n=187). C: Attitudes towards social prescribing (n=775).

Multiple demographic characteristics were associated with whether students had heard of SP (Figure 2; Table S2). Those who were older, studying at postgraduate levels (masters/doctoral vs undergraduate), and studying natural sciences or professions/applied sciences (vs social science) had higher odds of being aware of SP. In contrast, students whose first language was not English and those of Asian/Asian British ethnicity (vs White) had lower odds of knowing about SP. Looking at health-related factors, those who reported mental or physical health difficulties had higher odds of being aware of SP, as did those with better mental wellbeing. Health behaviours associated with SP awareness included speaking to a professional about mental health, engaging in social or community activities, and missing work due to mental health, all of which increased the odds of knowing about SP.

**Figure 2.**
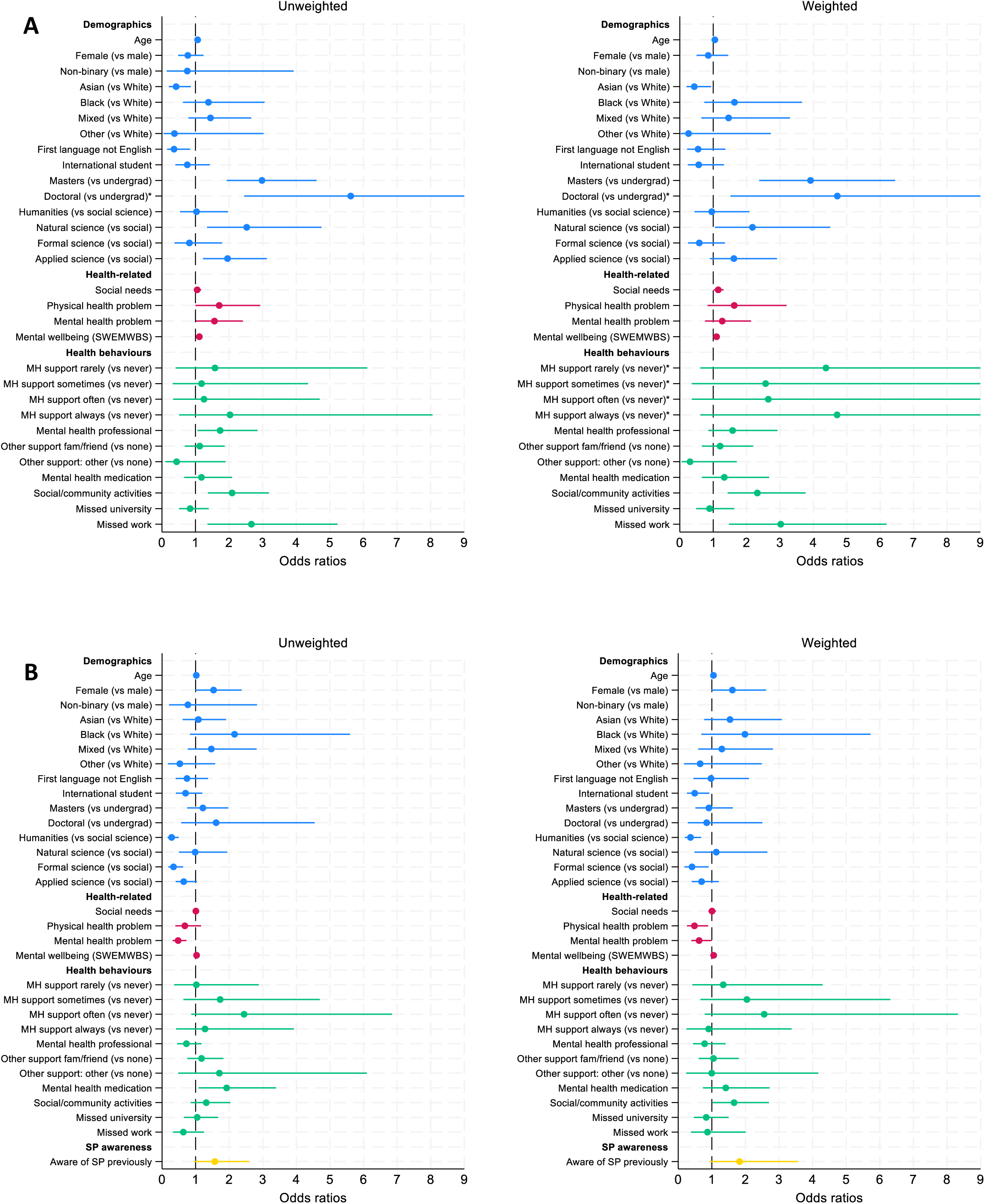
Factors associated with A) awareness of social prescribing and B) positive attitudes towards social prescribing (unweighted n=775, weighted n=761). Demographic, health-related, health behaviour, and SP awareness tested in sequential models to address confounding. *Odds ratios and 95% confidence intervals shown, but 95% confidence intervals are truncated for variables indicated with asterisks due to extreme values (see Tables S2 & S4 for full results).

After weighting, results were qualitatively similar, although confidence intervals were wider (Table S4). Associations with age, ethnicity, level of study, mental wellbeing, engaging in activities, and missing work were replicated. Yet, there was no longer evidence that participants studying professions/applied sciences had higher odds of being aware of SP than those studying social science. Instead of associations with physical or mental health difficulties, there was evidence that those with more social needs had higher odds of having heard of SP previously. Finally, although the OR was similar, there was no longer evidence that speaking to a professional about mental health was associated with SP awareness.

### Attitudes towards social prescribing

Despite the lack of awareness of SP, attitudes towards SP were highly favourable following presentation of a standardised definition (Figure 1). Nearly all students (97%) believed SP could support mental health and wellbeing, 95% believed universities should offer SP, and 89% reported they would accept SP if offered by a healthcare professional. Overall, 92% agreed that SP is relevant to future health professionals. The attitude least often endorsed was that SP should be part of the curriculum for health sciences programmes, but 86% did still agree with this statement.

Using our dichotomised indicator of attitudes towards SP, 76% of participants had entirely positive attitudes. In contrast to awareness of SP, fewer factors were associated with attitudes towards to SP (Figure 2; Table S3). For demographics, there was only an association with academic field, as those studying humanities and maths/computing (formal sciences) had lower odds of positive attitudes than those studying social sciences. Looking at health-related factors, those with mental health problems had lower odds of positive attitudes towards SP. In contrast, students who had taken medication for their mental health were more likely to have positive attitudes towards SP. No other factors were associated with attitudes, including previous awareness of SP.

After weighting, results were again similar, although showed some further evidence for associations (Table S5). Associations with academic field and mental health problems were replicated. Additionally, students who were older had slightly higher odds of positive attitudes towards SP, whereas international students had lower odds than home students. Furthermore, those with physical health problems had lower odds of positive attitudes towards SP. Finally, looking at health behaviours, instead of an association with taking medication for mental health, students who had engaged in social or community activities had marginally higher odds of positive attitudes.

## Discussion

Most UK university students had never heard of SP. Only 25% were aware of SP, substantially lower than the 75% of medical students at UK universities with SP Student Champions in 2020,^30^ although still much higher than the 7% of medical students who had heard of SP in 2017/2018.^29^ Our findings also show more awareness of SP in the UK than in a Canadian university in 2023, where only 16% of students had heard of SP.^31^ Together, these comparisons demonstrate the importance of measuring awareness amongst students across all disciplines. Findings from medical students cannot be generalised to the wider student body, and awareness of SP may differ across countries. Given that the SP movement began in the UK, before expanding to over 30 countries globally,^41^ it is not surprising the awareness was higher in the UK than in Canada. However, the difference to the Canadian sample could also be a result of awareness increasing over time as well as SP becoming more likely to be mentioned in courses, particularly within health and social sciences.

We also extended previous research by describing how awareness of and attitudes towards SP differed across sociodemographic groups for the first time. Given that SP has been proposed to address health inequalities,^42,43^ it was concerning that students whose first language was not English and those of Asian/Asian British ethnicity were less likely to have heard about SP. This could indicate that students facing more structural disadvantages may be least likely to access SP, meaning it risks widening inequalities rather than addressing them. However, in weighted analyses, those with more social needs were more likely to have heard of SP, which is in line with previous findings that people living in more deprived areas are increasingly likely to have been referred to SP.^21,44^ It was not surprising that students with health difficulties were more likely to be aware of SP as they may have found out about it through searching online or seeking advice from others. But we did not expect those with better wellbeing to also be more likely to have heard of SP. It could be that these students have better wellbeing because they are more aware of the importance of lifestyle, health behaviours, and care pathways such as SP.

As in previous research, we found an awareness-attitude paradox.^26,29–31^ Although only a quarter of students had heard of SP, attitudes towards SP were overwhelmingly positive once the concept was explained. Overall, 97% agreed that SP could support mental health and wellbeing, 95% felt that their university should offer SP, and 89% were willing to follow a healthcare professional’s advice if they were to suggest SP. These extremely high rates of agreement suggest that, if SP were to be implemented in UK universities, there could be very high uptake. The principal challenge for implementing university social prescribing may not be persuading students of its value but ensuring that they know the pathway exists and understand how to access it. In contrast to awareness of SP, very few factors were associated with positive attitudes towards SP. This is likely because students rated SP so favourably, with only 24% not reporting entirely positive attitudes. Although awareness of SP was not associated with attitudes, academic field was important, with those studying social sciences more likely to have positive attitudes. This suggests that education about health and healthcare (as is common in the social sciences) may encourage students to think more positively about SP.

Our findings have several clear implications for the development of university SP. Communication around and promotion of SP schemes will be key for ensuring uptake, as lack of awareness about what SP involves is likely to be a key barrier. Previous studies have shown that teaching can increase knowledge of SP,^26,29,30^ but it may be particularly important to target outreach to specific groups who are less likely to have heard of SP or have fewer positive attitudes towards SP. In the UK, this could include students who are younger, of Asian/Asian British ethnicity, have a first language other than English, not studying health-related subjects, and students with lower wellbeing or health problems.

We have also identified a clear gap for SP, as only around a third of students reported doing social or community activities to support their health and wellbeing in the last three months. Among these, a small minority did so through SP. So, although some students are already engaged in the type of activities they might be referred to via SP, the majority are not. This highlights a missed opportunity to formalise referral pathways and connect students to existing resources, both within the university and the wider community, through SP. Based on the positive attitudes shown in this study, SP may be better suited to the increasingly diverse student population and less stigmatising than other university mental health services, reducing common barriers to help-seeking.^11,25^ Universities also present unique opportunities for peer delivery of SP, as students could themselves be trained as link workers, embedding this pathway as a coproduced model and providing learning and experience in health and social care.^25^

This study has several strengths, including inclusion of students from numerous academic fields and stages of study at universities across the UK. It was the first UK study to explore student awareness and perceptions of SP in the wider student body, not just medical students. Measures of SP awareness and attitudes were designed following previous approaches,^29,31^ making our findings comparable across contexts. However, the binary measures of attitudes towards SP did result in little variation, so could be more detailed in future research. We also relied on a convenient sample who responded to adverts shared on social media and via email, meaning the sample could be biased towards those interested in SP. This could explain why those studying social sciences and professions/applied sciences were overrepresented. Although probability weights made the sample more representative of UK university students, academic field could not be included in the weight generation as it led to unstable estimates. There were some inconsistencies between unweighted and weighted analyses, which indicates that some of the unweighted findings may have been driven by over-represented groups. It will be important for future research on university SP to use more robust approaches to sampling to include larger and more representative samples. Finally, although the survey and findings were discussed with university students, this was not a student-led study. Future research should include student co-researchers to ensure it is coproduced.

UK university student numbers are increasing but the health and wellbeing of this population is declining. Among a field that generally focusses on student mental health problems,^45^ SP offers a positive person-centred approach that could be peer led.^25,26^ Although three quarters of students were not yet aware of SP, their positive attitudes towards SP suggest it could be well-suited to address the complex challenges that students face. The combination of low awareness and high acceptability suggests that implementation efforts should prioritise awareness-raising and clear referral pathways, rather than attempting to increase students’ willingness to engage with SP itself. Students, charities, and educators have emphasized the need for whole university approaches to support student health and wellbeing.^11,24,25,46^ SP fits perfectly into this framework, connecting students into existing resources across the university and community. We therefore recommend that UK universities consider implementing SP pathways, alongside developing communication and promotion materials for SP, both for future healthcare professionals and the wider student body.

## Data Availability

These data cannot be shared publicly. Code for the analyses is available from JKB on request.

## Declarations

### Author contributions

DH conceptualised the study and was responsible for the data collection. DH and JKB developed the analytical plan, with input from DF. JKB drafted the manuscript. All co-authors contributed to the interpretation and reporting of findings and read and approved the final version of the manuscript.

### Data sharing

These data cannot be shared publicly. Code for the analyses is available from JKB on request.

## Supplementary Materials

### Definition of social prescribing provided in the survey

An individual has a non-medical, health related social need. They are referred through to a ‘link worker’ or ‘social prescriber’ who spends time with the individual to understand their needs, strengths and interests. From this, they develop a personalised action plan to connect the individual to community resources and facilitate the individual accessing identified sources of support. They check in with the individual after they have attended to see how things went and if the sources of support if right for them. Example: A student visits student health services and indicates that they are feeling lonely. They are connected to a link worker, who is able to help address their needs and connect them with services, such as a new student society or a community group based on the students’ interests.

**Table S1.**
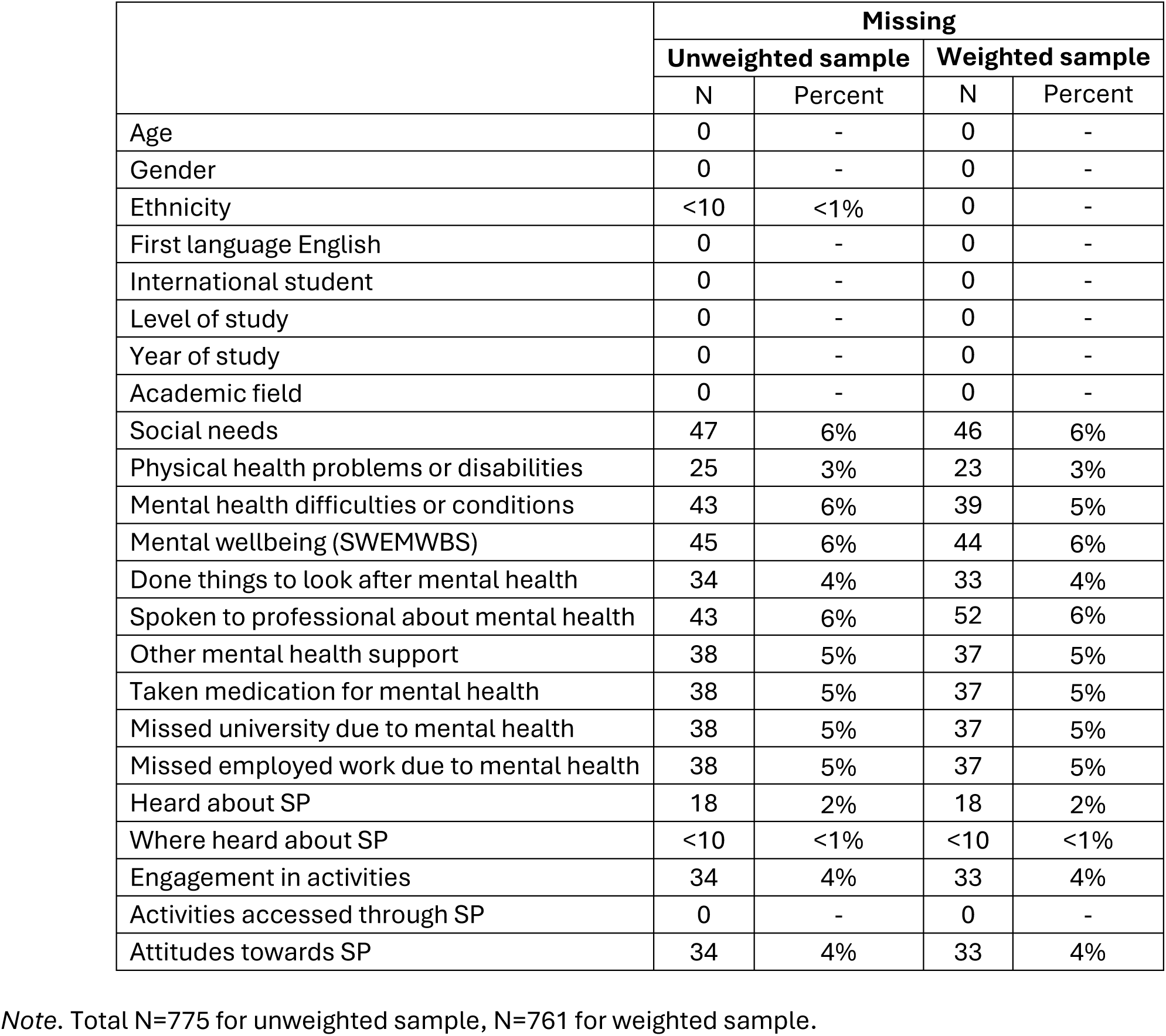
Missingness on included variables before limiting sample to complete cases.

|  | Missing |  |  |  |
| --- | --- | --- | --- | --- |
|  | Unweighted sample |  | Weighted sample |  |
|  | N | Percent | N | Percent |
| Age | 0 | - | 0 | - |
| Gender | 0 | - | 0 | - |
| Ethnicity | <10 | <1% | 0 | - |
| First language English | 0 | - | 0 | - |
| International student | 0 | - | 0 | - |
| Level of study | 0 | - | 0 | - |
| Year of study | 0 | - | 0 | - |
| Academic field | 0 | - | 0 | - |
| Social needs | 47 | 6% | 46 | 6% |
| Physical health problems or disabilities | 25 | 3% | 23 | 3% |
| Mental health difficulties or conditions | 43 | 6% | 39 | 5% |
| Mental wellbeing (SWEMWBS) | 45 | 6% | 44 | 6% |
| Done things to look after mental health | 34 | 4% | 33 | 4% |
| Spoken to professional about mental health | 43 | 6% | 52 | 6% |
| Other mental health support | 38 | 5% | 37 | 5% |
| Taken medication for mental health | 38 | 5% | 37 | 5% |
| Missed university due to mental health | 38 | 5% | 37 | 5% |
| Missed employed work due to mental health | 38 | 5% | 37 | 5% |
| Heard about SP | 18 | 2% | 18 | 2% |
| Where heard about SP | <10 | <1% | <10 | <1% |
| Engagement in activities | 34 | 4% | 33 | 4% |
| Activities accessed through SP | 0 | - | 0 | - |
| Attitudes towards SP | 34 | 4% | 33 | 4% |

Note. Total N=775 for unweighted sample, N=761 for weighted sample.

### Imputed analyses (unweighted)

**Table S2.** Associations between individual characteristics and awareness of social prescribing.

|  | Model 1 |  |  | Model 2 |  |  | Model 3 |  |  |
| --- | --- | --- | --- | --- | --- | --- | --- | --- | --- |
|  | OR | 95% CI | p | OR | 95% CI | p | OR | 95% CI | p |
| Age | **1.06** | **1.04, 1.09** | **<0.001** | 1.05 | 1.03, 1.08 | <0.001 | 1.06 | 1.03, 1.08 | <0.001 |
| Gender (ref: male) |  |  |  |  |  |  |  |  |  |
| Female | 0.77 | 0.48, 1.24 | 0.282 | 0.77 | 0.47, 1.25 | 0.284 | 0.75 | 0.45, 1.25 | 0.265 |
| Non-binary/other | 0.76 | 0.15, 3.92 | 0.739 | 0.73 | 0.13, 4.18 | 0.722 | 0.53 | 0.08, 3.34 | 0.501 |
| Ethnicity (ref: White) |  |  |  |  |  |  |  |  |  |
| Asian/Asian British | **0.42** | **0.21, 0.86** | **0.017** | 0.44 | 0.21, 0.90 | 0.026 | 0.61 | 0.29, 1.28 | 0.190 |
| Black/African/Caribbean/Black British | 1.39 | 0.63, 3.06 | 0.420 | 1.34 | 0.59, 3.05 | 0.490 | 1.10 | 0.44, 2.76 | 0.844 |
| Mixed/multiple ethnic groups | 1.45 | 0.79, 2.66 | 0.235 | 1.16 | 0.60, 2.24 | 0.663 | 0.91 | 0.45, 1.85 | 0.795 |
| Other | 0.37 | 0.05, 3.03 | 0.357 | 0.34 | 0.04, 2.84 | 0.321 | 0.31 | 0.04, 2.63 | 0.280 |
| First language not English (ref: English) | **0.36** | **0.15, 0.84** | **0.018** | 0.44 | 0.19, 1.04 | 0.062 | 0.43 | 0.18, 1.05 | 0.063 |
| International student (ref: home student) | 0.76 | 0.40, 1.43 | 0.386 | 0.51 | 0.26, 1.00 | 0.050 | 0.44 | 0.22, 0.91 | 0.026 |
| Level of study (ref: undergraduate) |  |  |  |  |  |  |  |  |  |
| Masters | **2.98** | **1.93, 4.60** | **<0.001** | 3.44 | 2.18, 5.45 | <0.001 | 3.35 | 2.08, 5.39 | <0.001 |
| Doctoral | **5.62** | **2.45, 12.90** | **<0.001** | 5.39 | 2.25, 12.90 | <0.001 | 5.02 | 2.05, 12.25 | <0.001 |
| Academic field (ref: social science) |  |  |  |  |  |  |  |  |  |
| Humanities | 1.03 | 0.54, 1.97 | 0.927 | 1.00 | 0.51, 1.93 | 0.993 | 1.03 | 0.51, 2.09 | 0.930 |
| Natural sciences | **2.53** | **1.34, 4.75** | **0.004** | 2.56 | 1.33, 4.91 | 0.005 | 2.59 | 1.31, 5.13 | 0.006 |
| Maths/computing (formal science) | 0.82 | 0.37, 1.79 | 0.616 | 0.87 | 0.39, 1.95 | 0.731 | 1.02 | 0.44, 2.36 | 0.972 |
| Professions and applied sciences | **1.95** | **1.22, 3.12** | **0.005** | 2.17 | 1.34, 3.53 | 0.002 | 2.20 | 1.34, 3.64 | 0.002 |
| Social needs | - | - | - | 1.05 | 0.95, 1.16 | 0.351 | 1.02 | 0.91, 1.14 | 0.726 |
| Physical health problems or disabilities | - | - | - | **1.71** | **0.99, 2.93** | **0.052** | 1.84 | 1.04, 3.25 | 0.037 |
| Mental health difficulties or conditions | - | - | - | **1.56** | **1.02, 2.41** | **0.042** | 1.10 | 0.63, 1.91 | 0.743 |
| Mental wellbeing (SWEMWBS) | - | - | - | **1.11** | **1.06, 1.17** | **<0.001** | 1.11 | 1.05, 1.17 | <0.001 |
| Done things to look after mental health (ref: never) |  |  |  |  |  |  |  |  |  |
| Rarely | - | - | - | - | - | - | 1.58 | 0.41, 6.12 | 0.507 |
| Sometimes | - | - | - | - | - | - | 1.18 | 0.32, 4.35 | 0.803 |
| Often | - | - | - | - | - | - | 1.25 | 0.33, 4.70 | 0.737 |
| Always | - | - | - | - | - | - | 2.03 | 0.51, 8.06 | 0.315 |
| Spoken to professional about mental health | - | - | - | - | - | - | **1.73** | **1.06, 2.85** | **0.029** |
| Other mental health support (ref: none) |  |  |  |  |  |  |  |  |  |
| Family/friend | - | - | - | - | - | - | 1.13 | 0.68, 1.87 | 0.643 |
| Other | - | - | - | - | - | - | 0.44 | 0.10, 1.89 | 0.269 |
| Taken medication for mental health | - | - | - | - | - | - | 1.18 | 0.66, 2.09 | 0.578 |
| Engaged in social/community activities | - | - | - | - | - | - | **2.09** | **1.37, 3.19** | **0.001** |
| Missed university due to mental health | - | - | - | - | - | - | 0.84 | 0.51, 1.39 | 0.501 |
| Missed employed work due to mental health | - | - | - | - | - | - | **2.66** | **1.36, 5.22** | **0.004** |

Note. N=775. OR: odds ratio. Bold text indicates p<0.05. Findings interpreted from Model 1 for sociodemographics, Model 2 for health-related factors, and Model 3 for health behaviours.

**Table S3.** Associations between individual characteristics and overall attitudes towards social prescribing.

|  | Model 1 |  |  | Model 2 |  |  | Model 3 |  |  | Model 4 |  |  |
| --- | --- | --- | --- | --- | --- | --- | --- | --- | --- | --- | --- | --- |
|  | OR | 95% CI | p | OR | 95% CI | p | OR | 95% CI | p | OR | 95% CI | p |
| Age | 1.02 | 0.99, 1.05 | 0.160 | 1.03 | 1.00, 1.06 | 0.051 | 1.03 | 1.00, 1.07 | 0.035 | 1.03 | 1.00, 1.06 | 0.065 |
| Gender (ref: male) |  |  |  |  |  |  |  |  |  |  |  |  |
| Female | 1.53 | 0.99, 2.37 | 0.054 | 1.69 | 1.08, 2.65 | 0.022 | 1.63 | 1.02, 2.61 | 0.041 | 1.65 | 1.03, 2.65 | 0.036 |
| Non-binary/other | 0.77 | 0.21, 2.83 | 0.691 | 1.09 | 0.29, 4.06 | 0.896 | 0.75 | 0.19, 2.96 | 0.679 | 0.77 | 0.19, 3.09 | 0.717 |
| Ethnicity (ref: White) |  |  |  |  |  |  |  |  |  |  |  |  |
| Asian/Asian British | 1.08 | 0.62, 1.90 | 0.782 | 0.85 | 0.48, 1.51 | 0.578 | 0.89 | 0.49, 1.62 | 0.714 | 0.91 | 0.50, 1.65 | 0.753 |
| Black/African/Caribbean/Black British | 2.16 | 0.83, 5.60 | 0.113 | 1.69 | 0.62, 4.56 | 0.303 | 1.94 | 0.70, 5.37 | 0.199 | 1.90 | 0.69, 5.24 | 0.216 |
| Mixed/multiple ethnic groups | 1.47 | 0.76, 2.81 | 0.250 | 1.53 | 0.76, 3.07 | 0.229 | 1.63 | 0.80, 3.30 | 0.177 | 1.63 | 0.80, 3.31 | 0.181 |
| Other | 0.53 | 0.18, 1.58 | 0.254 | 0.38 | 0.13, 1.18 | 0.094 | 0.41 | 0.13, 1.30 | 0.129 | 0.43 | 0.13, 1.38 | 0.155 |
| First language not English (ref: English) | 0.74 | 0.40, 1.37 | 0.339 | 0.64 | 0.34, 1.19 | 0.160 | 0.71 | 0.37, 1.35 | 0.293 | 0.73 | 0.38, 1.39 | 0.336 |
| International student (ref: home student) | 0.70 | 0.41, 1.20 | 0.191 | 0.83 | 0.47, 1.45 | 0.505 | 0.82 | 0.45, 1.48 | 0.503 | 0.86 | 0.47, 1.57 | 0.618 |
| Level of study (ref: undergraduate) |  |  |  |  |  |  |  |  |  |  |  |  |
| Masters | 1.21 | 0.75, 1.97 | 0.429 | 1.24 | 0.76, 2.03 | 0.395 | 1.13 | 0.68, 1.88 | 0.641 | 1.04 | 0.62, 1.75 | 0.871 |
| Doctoral | 1.61 | 0.57, 4.54 | 0.369 | 1.61 | 0.56, 4.66 | 0.379 | 1.56 | 0.53, 4.56 | 0.420 | 1.33 | 0.45, 3.96 | 0.603 |
| Academic field (ref: social science) |  |  |  |  |  |  |  |  |  |  |  |  |
| Humanities | **0.29** | **0.16, 0.50** | **<0.001** | 0.30 | 0.17, 0.52 | **<0.001** | 0.33 | 0.18, 0.61 | **<0.001** | 0.34 | 0.18, 0.61 | **<0.001** |
| Natural sciences | 0.99 | 0.50, 1.93 | 0.966 | 0.97 | 0.49, 1.92 | 0.923 | 1.02 | 0.51, 2.06 | 0.950 | 0.96 | 0.48, 1.95 | 0.919 |
| Maths/computing (formal science) | **0.34** | **0.19, 0.63** | **0.001** | 0.32 | 0.17, 0.60 | **<0.001** | 0.36 | 0.19, 0.68 | 0.002 | 0.36 | 0.19, 0.68 | 0.002 |
| Professions and applied sciences | 0.64 | 0.40, 1.03 | 0.069 | 0.62 | 0.39, 1.01 | 0.054 | 0.60 | 0.37, 0.98 | 0.042 | 0.57 | 0.35, 0.93 | 0.025 |
| Social needs | - | - | - | 1.01 | 0.92, 1.11 | 0.881 | 1.02 | 0.92, 1.12 | 0.735 | 1.02 | 0.92, 1.13 | 0.721 |
| Physical health problems or disabilities | - | - | - | 0.68 | 0.40, 1.16 | 0.157 | 0.77 | 0.44, 1.34 | 0.356 | 0.73 | 0.42, 1.29 | 0.282 |
| Mental health difficulties or conditions | - | - | - | **0.48** | **0.32, 0.72** | **<0.001** | 0.38 | 0.22, 0.64 | **<0.001** | 0.38 | 0.22, 0.65 | **<0.001** |
| Mental wellbeing (SWEMWBS) | - | - | - | 1.03 | 0.98, 1.08 | 0.265 | 1.02 | 0.96, 1.07 | 0.544 | 1.01 | 0.96, 1.07 | 0.679 |
| Done things to look after mental health (ref: never) |  |  |  |  |  |  |  |  |  |  |  |  |
| Rarely | - | - | - | - | - | - | 1.02 | 0.36, 2.88 | 0.968 | 0.99 | 0.35, 2.81 | 0.985 |
| Sometimes | - | - | - | - | - | - | 1.73 | 0.64, 4.69 | 0.281 | 1.69 | 0.62, 4.62 | 0.303 |
| Often | - | - | - | - | - | - | 2.44 | 0.87, 6.85 | 0.090 | 2.40 | 0.85, 6.78 | 0.098 |
| Always | - | - | - | - | - | - | 1.28 | 0.42, 3.93 | 0.667 | 1.22 | 0.39, 3.77 | 0.734 |
| Spoken to professional about mental health | - | - | - | - | - | - | 0.72 | 0.45, 1.18 | 0.191 | 0.69 | 0.43, 1.13 | 0.143 |
| Other mental health support (ref: none) |  |  |  |  |  |  |  |  |  |  |  |  |
| Family/friend | - | - | - | - | - | - | 1.17 | 0.75, 1.83 | 0.476 | 1.18 | 0.76, 1.84 | 0.465 |
| Other | - | - | - | - | - | - | 1.71 | 0.48, 6.10 | 0.410 | 1.77 | 0.50, 6.30 | 0.375 |
| Taken medication for mental health | - | - | - | - | - | - | **1.92** | **1.09, 3.39** | **0.024** | 1.89 | 1.07, 3.35 | 0.028 |
| Engaged in social/community activities | - | - | - | - | - | - | 1.32 | 0.85, 2.03 | 0.213 | 1.26 | 0.81, 1.94 | 0.302 |
| Missed university due to mental health | - | - | - | - | - | - | 1.05 | 0.66, 1.67 | 0.847 | 1.05 | 0.66, 1.68 | 0.827 |
| Missed employed work due to mental health | - | - | - | - | - | - | 0.63 | 0.32, 1.25 | 0.186 | 0.59 | 0.30, 1.16 | 0.126 |
| Aware of SP previously | - | - | - | - | - | - | - | - | - | 1.57 | 0.95, 2.60 | 0.078 |

Note. N=775. OR: odds ratio. Bold text indicates p<0.05. Findings interpreted from Model 1 for sociodemographics, Model 2 for health-related factors, and Model 3 for health behaviours.

### Weighted imputed analyses

**Table S4.** Associations between individual characteristics and awareness of social prescribing.

|  | Model 1 |  |  | Model 2 |  |  | Model 3 |  |  |
| --- | --- | --- | --- | --- | --- | --- | --- | --- | --- |
|  | OR | 95% CI | p | OR | 95% CI | p | OR | 95% CI | p |
| Age | **1.05** | **1.02, 1.08** | **0.003** | 1.04 | 1.00, 1.08 | 0.033 | 1.05 | 1.02, 1.08 | 0.001 |
| Gender (ref: male) |  |  |  |  |  |  |  |  |  |
| Female | 0.85 | 0.50, 1.45 | 0.553 | 0.94 | 0.55, 1.61 | 0.824 | 0.86 | 0.48, 1.54 | 0.608 |
| Non-binary/other | - | - | - | - | - | - | - | - | - |
| Ethnicity (ref: White) |  |  |  |  |  |  |  |  |  |
| Asian/Asian British | **0.43** | **0.20, 0.94** | **0.033** | 0.46 | 0.21, 1.00 | 0.051 | 0.65 | 0.28, 1.51 | 0.313 |
| Black/African/Caribbean/Black British | 1.64 | 0.73, 3.66 | 0.229 | 1.33 | 0.59, 3.01 | 0.487 | 0.88 | 0.37, 2.08 | 0.769 |
| Mixed/multiple ethnic groups | 1.46 | 0.65, 3.30 | 0.362 | 1.04 | 0.41, 2.62 | 0.934 | 0.75 | 0.28, 1.98 | 0.555 |
| Other | 0.26 | 0.02, 2.73 | 0.261 | 0.22 | 0.02, 2.46 | 0.218 | 0.21 | 0.02, 2.55 | 0.218 |
| First language not English (ref: English) | 0.55 | 0.22, 1.37 | 0.196 | 0.67 | 0.26, 1.68 | 0.390 | 0.66 | 0.27, 1.61 | 0.364 |
| International student (ref: home student) | 0.57 | 0.24, 1.32 | 0.186 | 0.37 | 0.16, 0.85 | 0.019 | 0.34 | 0.14, 0.80 | 0.014 |
| Level of study (ref: undergraduate) |  |  |  |  |  |  |  |  |  |
| Masters | **3.91** | **2.38, 6.45** | **<0.001** | 4.97 | 2.96, 8.32 | **<0.001** | 5.00 | 2.89, 8.65 | **<0.001** |
| Doctoral | **4.72** | **1.52, 14.69** | **0.007** | 5.98 | 1.94, 18.46 | 0.002 | 5.34 | 1.64, 17.40 | 0.006 |
| Academic field (ref: social science) |  |  |  |  |  |  |  |  |  |
| Humanities | 0.96 | 0.44, 2.08 | 0.912 | 0.90 | 0.40, 2.03 | 0.807 | 0.83 | 0.35, 1.93 | 0.660 |
| Natural sciences | **2.18** | **1.05, 4.50** | **0.036** | 2.14 | 1.01, 4.52 | 0.046 | 2.09 | 0.92, 4.75 | 0.077 |
| Maths/computing (formal science) | 0.58 | 0.25, 1.35 | 0.208 | 0.53 | 0.22, 1.26 | 0.151 | 0.65 | 0.26, 1.62 | 0.353 |
| Professions and applied sciences | 1.62 | 0.90, 2.91 | 0.107 | 1.77 | 0.98, 3.19 | 0.058 | 1.70 | 0.93, 3.09 | 0.083 |
| Social needs | - | - | - | **1.15** | **1.01, 1.31** | **0.032** | 1.11 | 0.97, 1.27 | 0.113 |
| Physical health problems or disabilities | - | - | - | 1.63 | 0.83, 3.19 | 0.154 | 1.85 | 0.93, 3.70 | 0.081 |
| Mental health difficulties or conditions | - | - | - | 1.27 | 0.76, 2.13 | 0.366 | 0.85 | 0.43, 1.70 | 0.645 |
| Mental wellbeing (SWEMWBS) | - | - | - | **1.10** | **1.03, 1.17** | **0.004** | 1.09 | 1.02, 1.16 | 0.010 |
| Done things to look after mental health (ref: never) |  |  |  |  |  |  |  |  |  |
| Rarely | - | - | - | - | - | - | 4.38 | 0.62, 31.11 | 0.140 |
| Sometimes | - | - | - | - | - | - | 2.57 | 0.36, 18.47 | 0.348 |
| Often | - | - | - | - | - | - | 2.65 | 0.36, 19.44 | 0.337 |
| Always | - | - | - | - | - | - | 4.71 | 0.61, 36.17 | 0.136 |
| Spoken to professional about mental health | - | - | - | - | - | - | 1.58 | 0.86, 2.92 | 0.143 |
| Other mental health support (ref: none) |  |  |  |  |  |  |  |  |  |
| Family/friend | - | - | - | - | - | - | 1.21 | 0.66, 2.20 | 0.537 |
| Other | - | - | - | - | - | - | 0.31 | 0.06, 1.70 | 0.176 |
| Taken medication for mental health | - | - | - | - | - | - | 1.33 | 0.66, 2.67 | 0.419 |
| Engaged in social/community activities | - | - | - | - | - | - | **2.32** | **1.43, 3.76** | **0.001** |
| Missed university due to mental health | - | - | - | - | - | - | 0.89 | 0.49, 1.63 | 0.710 |
| Missed employed work due to mental health | - | - | - | - | - | - | **3.02** | **1.47, 6.19** | **0.003** |

Note. N=761. OR: odds ratio. Bold text indicates p<0.05. Findings interpreted from Model 1 for sociodemographics, Model 2 for health-related factors, and Model 3 for health behaviours.

**Table S5.**
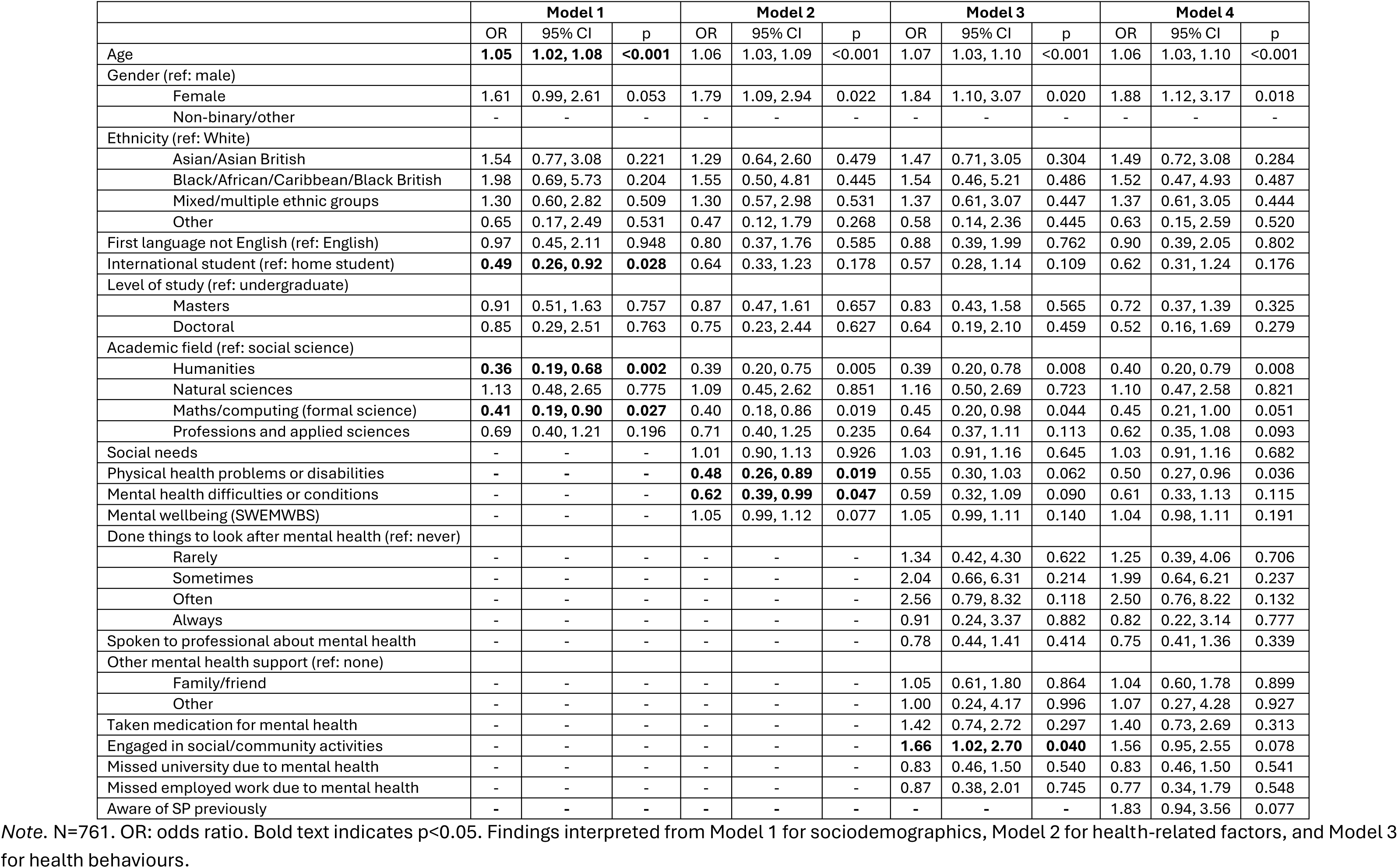
Associations between individual characteristics and overall attitudes towards social prescribing.

|  | Model 1 |  |  | Model 2 |  |  | Model 3 |  |  | Model 4 |  |  |
| --- | --- | --- | --- | --- | --- | --- | --- | --- | --- | --- | --- | --- |
|  | OR | 95% CI | p | OR | 95% CI | p | OR | 95% CI | p | OR | 95% CI | p |
| Age | **1.05** | **1.02, 1.08** | **<0.001** | 1.06 | 1.03, 1.09 | <0.001 | 1.07 | 1.03, 1.10 | <0.001 | 1.06 | 1.03, 1.10 | <0.001 |
| Gender (ref: male) |  |  |  |  |  |  |  |  |  |  |  |  |
| Female | 1.61 | 0.99, 2.61 | 0.053 | 1.79 | 1.09, 2.94 | 0.022 | 1.84 | 1.10, 3.07 | 0.020 | 1.88 | 1.12, 3.17 | 0.018 |
| Non-binary/other | - | - | - | - | - | - | - | - | - | - | - | - |
| Ethnicity (ref: White) |  |  |  |  |  |  |  |  |  |  |  |  |
| Asian/Asian British | 1.54 | 0.77, 3.08 | 0.221 | 1.29 | 0.64, 2.60 | 0.479 | 1.47 | 0.71, 3.05 | 0.304 | 1.49 | 0.72, 3.08 | 0.284 |
| Black/African/Caribbean/Black British | 1.98 | 0.69, 5.73 | 0.204 | 1.55 | 0.50, 4.81 | 0.445 | 1.54 | 0.46, 5.21 | 0.486 | 1.52 | 0.47, 4.93 | 0.487 |
| Mixed/multiple ethnic groups | 1.30 | 0.60, 2.82 | 0.509 | 1.30 | 0.57, 2.98 | 0.531 | 1.37 | 0.61, 3.07 | 0.447 | 1.37 | 0.61, 3.05 | 0.444 |
| Other | 0.65 | 0.17, 2.49 | 0.531 | 0.47 | 0.12, 1.79 | 0.268 | 0.58 | 0.14, 2.36 | 0.445 | 0.63 | 0.15, 2.59 | 0.520 |
| First language not English (ref: English) | 0.97 | 0.45, 2.11 | 0.948 | 0.80 | 0.37, 1.76 | 0.585 | 0.88 | 0.39, 1.99 | 0.762 | 0.90 | 0.39, 2.05 | 0.802 |
| International student (ref: home student) | **0.49** | **0.26, 0.92** | **0.028** | 0.64 | 0.33, 1.23 | 0.178 | 0.57 | 0.28, 1.14 | 0.109 | 0.62 | 0.31, 1.24 | 0.176 |
| Level of study (ref: undergraduate) |  |  |  |  |  |  |  |  |  |  |  |  |
| Masters | 0.91 | 0.51, 1.63 | 0.757 | 0.87 | 0.47, 1.61 | 0.657 | 0.83 | 0.43, 1.58 | 0.565 | 0.72 | 0.37, 1.39 | 0.325 |
| Doctoral | 0.85 | 0.29, 2.51 | 0.763 | 0.75 | 0.23, 2.44 | 0.627 | 0.64 | 0.19, 2.10 | 0.459 | 0.52 | 0.16, 1.69 | 0.279 |
| Academic field (ref: social science) |  |  |  |  |  |  |  |  |  |  |  |  |
| Humanities | **0.36** | **0.19, 0.68** | **0.002** | 0.39 | 0.20, 0.75 | 0.005 | 0.39 | 0.20, 0.78 | 0.008 | 0.40 | 0.20, 0.79 | 0.008 |
| Natural sciences | 1.13 | 0.48, 2.65 | 0.775 | 1.09 | 0.45, 2.62 | 0.851 | 1.16 | 0.50, 2.69 | 0.723 | 1.10 | 0.47, 2.58 | 0.821 |
| Maths/computing (formal science) | **0.41** | **0.19, 0.90** | **0.027** | 0.40 | 0.18, 0.86 | 0.019 | 0.45 | 0.20, 0.98 | 0.044 | 0.45 | 0.21, 1.00 | 0.051 |
| Professions and applied sciences | 0.69 | 0.40, 1.21 | 0.196 | 0.71 | 0.40, 1.25 | 0.235 | 0.64 | 0.37, 1.11 | 0.113 | 0.62 | 0.35, 1.08 | 0.093 |
| Social needs | - | - | - | 1.01 | 0.90, 1.13 | 0.926 | 1.03 | 0.91, 1.16 | 0.645 | 1.03 | 0.91, 1.16 | 0.682 |
| Physical health problems or disabilities | - | - | - | **0.48** | **0.26, 0.89** | **0.019** | 0.55 | 0.30, 1.03 | 0.062 | 0.50 | 0.27, 0.96 | 0.036 |
| Mental health difficulties or conditions | - | - | - | **0.62** | **0.39, 0.99** | **0.047** | 0.59 | 0.32, 1.09 | 0.090 | 0.61 | 0.33, 1.13 | 0.115 |
| Mental wellbeing (SWEMWBS) | - | - | - | 1.05 | 0.99, 1.12 | 0.077 | 1.05 | 0.99, 1.11 | 0.140 | 1.04 | 0.98, 1.11 | 0.191 |
| Done things to look after mental health (ref: never) |  |  |  |  |  |  |  |  |  |  |  |  |
| Rarely | - | - | - | - | - | - | 1.34 | 0.42, 4.30 | 0.622 | 1.25 | 0.39, 4.06 | 0.706 |
| Sometimes | - | - | - | - | - | - | 2.04 | 0.66, 6.31 | 0.214 | 1.99 | 0.64, 6.21 | 0.237 |
| Often | - | - | - | - | - | - | 2.56 | 0.79, 8.32 | 0.118 | 2.50 | 0.76, 8.22 | 0.132 |
| Always | - | - | - | - | - | - | 0.91 | 0.24, 3.37 | 0.882 | 0.82 | 0.22, 3.14 | 0.777 |
| Spoken to professional about mental health | - | - | - | - | - | - | 0.78 | 0.44, 1.41 | 0.414 | 0.75 | 0.41, 1.36 | 0.339 |
| Other mental health support (ref: none) |  |  |  |  |  |  |  |  |  |  |  |  |
| Family/friend | - | - | - | - | - | - | 1.05 | 0.61, 1.80 | 0.864 | 1.04 | 0.60, 1.78 | 0.899 |
| Other | - | - | - | - | - | - | 1.00 | 0.24, 4.17 | 0.996 | 1.07 | 0.27, 4.28 | 0.927 |
| Taken medication for mental health | - | - | - | - | - | - | 1.42 | 0.74, 2.72 | 0.297 | 1.40 | 0.73, 2.69 | 0.313 |
| Engaged in social/community activities | - | - | - | - | - | - | **1.66** | **1.02, 2.70** | **0.040** | 1.56 | 0.95, 2.55 | 0.078 |
| Missed university due to mental health | - | - | - | - | - | - | 0.83 | 0.46, 1.50 | 0.540 | 0.83 | 0.46, 1.50 | 0.541 |
| Missed employed work due to mental health | - | - | - | - | - | - | 0.87 | 0.38, 2.01 | 0.745 | 0.77 | 0.34, 1.79 | 0.548 |
| Aware of SP previously | - | - | - | - | - | - | - | - | - | 1.83 | 0.94, 3.56 | 0.077 |

Note. N=761. OR: odds ratio. Bold text indicates p<0.05. Findings interpreted from Model 1 for sociodemographics, Model 2 for health-related factors, and Model 3 for health behaviours.

### Complete case analyses (unweighted)

**Table S6.** Sample characteristics.

|  | **Mean (SD)** |
| --- | --- |
| Age | 24.67 (7.49) |
| Social needs (range 0-9) | 2.61 (2.17) |
| Mental wellbeing (SWEMWBS, range 7-35) | 22.56 (4.35) |
|  | **Proportion** |
| Gender |  |
| Male | 21% |
| Female | 78% |
| Non-binary/other | 1% |
| Ethnicity |  |
| White | 65% |
| Asian/Asian British | 16% |
| Black/African/Caribbean/Black British | 6% |
| Mixed/multiple ethnic groups | 11% |
| Other | 2% |
| First language English | 88% |
| International student | 17% |
| Level of study |  |
| Undergraduate | 75% |
| Masters | 20% |
| Doctoral | 5% |
| Year of study |  |
| 1 | 37% |
| 2 | 24% |
| 3 | 23% |
| 4 | 12% |
| 5 | 2% |
| Other | 1% |
| Academic field |  |
| Social science | 39% |
| Humanities | 12% |
| Natural sciences | 11% |
| Maths/computing (formal science) | 9% |
| Professions and applied sciences | 30% |
| Physical health problems or disabilities | 17% |
| Mental health difficulties or conditions | 35% |
| Done things to look after mental health |  |
| Never | 3% |
| Rarely | 16% |
| Sometimes | 39% |
| Often | 31% |
| Always | 11% |
| Spoken to professional about mental health | 42% |
| Other mental health support |  |
| None | 33% |
| Family/friend | 64% |
| Other | 2% |
| Taken medication for mental health | 23% |
| Missed university due to mental health | 29% |
| Missed employed work due to mental health | 9% |

Note. N=692.

**Table S7.** Associations between individual characteristics and awareness of social prescribing.

|  | Model 1 |  |  | Model 2 |  |  | Model 3 |  |  |
| --- | --- | --- | --- | --- | --- | --- | --- | --- | --- |
|  | OR | 95% CI | p | OR | 95% CI | p | OR | 95% CI | p |
| Age | **1.07** | **1.05, 1.10** | **<0.001** | 1.06 | 1.03, 1.09 | <0.001 | 1.07 | 1.04, 1.10 | <0.001 |
| Gender (ref: male) |  |  |  |  |  |  |  |  |  |
| Female | 0.87 | 0.54, 1.41 | 0.576 | 0.84 | 0.51, 1.39 | 0.505 | 0.82 | 0.48, 1.38 | 0.456 |
| Non-binary/other | 1.38 | 0.25, 7.68 | 0.712 | 1.12 | 0.18, 6.78 | 0.904 | 0.83 | 0.12, 5.71 | 0.848 |
| Ethnicity (ref: White) |  |  |  |  |  |  |  |  |  |
| Asian/Asian British | **0.47** | **0.22, 1.00** | **0.049** | 0.49 | 0.23, 1.06 | 0.069 | 0.70 | 0.32, 1.53 | 0.369 |
| Black/African/Caribbean/Black British | 1.11 | 0.49, 2.48 | 0.808 | 1.14 | 0.49, 2.65 | 0.765 | 0.99 | 0.39, 2.53 | 0.986 |
| Mixed/multiple ethnic groups | 1.52 | 0.83, 2.80 | 0.178 | 1.17 | 0.59, 2.30 | 0.659 | 0.93 | 0.45, 1.92 | 0.841 |
| Other | 0.38 | 0.05, 3.04 | 0.360 | 0.38 | 0.05, 3.14 | 0.369 | 0.35 | 0.04, 3.04 | 0.338 |
| First language not English (ref: English) | **0.29** | **0.12, 0.70** | **0.006** | 0.36 | 0.14, 0.89 | 0.027 | 0.31 | 0.12, 0.81 | 0.016 |
| International student (ref: home student) | 0.89 | 0.49, 1.61 | 0.690 | 0.59 | 0.31, 1.14 | 0.116 | 0.52 | 0.26, 1.06 | 0.073 |
| Level of study (ref: undergraduate) |  |  |  |  |  |  |  |  |  |
| Masters | **3.18** | **2.04, 4.96** | **<0.001** | 3.50 | 2.20, 5.57 | <0.001 | 3.30 | 2.03, 5.35 | <0.001 |
| Doctoral | **4.42** | **1.88, 10.35** | **0.001** | 4.12 | 1.69, 10.02 | 0.002 | 4.03 | 1.63, 9.98 | 0.003 |
| Academic field (ref: social science) |  |  |  |  |  |  |  |  |  |
| Humanities | 0.99 | 0.52, 1.90 | 0.985 | 0.91 | 0.46, 1.77 | 0.774 | 0.87 | 0.42, 1.76 | 0.691 |
| Natural sciences | **2.66** | **1.41, 5.04** | **0.003** | 2.57 | 1.33, 4.96 | 0.005 | 2.47 | 1.25, 4.91 | 0.010 |
| Maths/computing (formal science) | 0.95 | 0.42, 2.12 | 0.898 | 0.95 | 0.42, 2.17 | 0.904 | 1.05 | 0.45, 2.49 | 0.904 |
| Professions and applied sciences | **1.81** | **1.11, 2.96** | **0.017** | 1.92 | 1.16, 3.18 | 0.011 | 1.95 | 1.16, 3.28 | 0.012 |
| Social needs | - | - | - | 1.05 | 0.94, 1.16 | 0.386 | 1.00 | 0.89, 1.12 | 0.969 |
| Physical health problems or disabilities | - | - | - | 1.52 | 0.88, 2.63 | 0.129 | 1.63 | 0.92, 2.90 | 0.096 |
| Mental health difficulties or conditions | - | - | - | **1.65** | **1.07, 2.56** | **0.024** | 1.16 | 0.67, 2.02 | 0.590 |
| Mental wellbeing (SWEMWBS) | - | - | - | **1.10** | **1.05, 1.15** | **<0.001** | 1.09 | 1.03, 1.15 | 0.002 |
| Done things to look after mental health (ref: never) |  |  |  |  |  |  |  |  |  |
| Rarely | - | - | - | - | - | - | 1.12 | 0.29, 4.32 | 0.864 |
| Sometimes | - | - | - | - | - | - | 0.87 | 0.24, 3.17 | 0.831 |
| Often | - | - | - | - | - | - | 0.90 | 0.25, 3.30 | 0.875 |
| Always | - | - | - | - | - | - | 1.33 | 0.34, 5.24 | 0.681 |
| Spoken to professional about mental health | - | - | - | - | - | - | **1.77** | **1.07, 2.95** | **0.027** |
| Other mental health support (ref: none) |  |  |  |  |  |  |  |  |  |
| Family/friend | - | - | - | - | - | - | 1.17 | 0.71, 1.94 | 0.540 |
| Other | - | - | - | - | - | - | 0.48 | 0.12, 1.93 | 0.302 |
| Taken medication for mental health | - | - | - | - | - | - | 1.08 | 0.61, 1.93 | 0.794 |
| Engaged in social/community activities | - | - | - | - | - | - | **2.14** | **1.39, 3.30** | **0.001** |
| Missed university due to mental health | - | - | - | - | - | - | 0.94 | 0.56, 1.57 | 0.802 |
| Missed employed work due to mental health | - | - | - | - | - | - | **2.92** | **1.43, 5.96** | **0.003** |

Note. N=692. OR: odds ratio. Bold text indicates p<0.05.

**Table S8.** Associations between individual characteristics and overall attitudes towards social prescribing.

|  | Model 1 |  |  | Model 2 |  |  | Model 3 |  |  | Model 4 |  |  |
| --- | --- | --- | --- | --- | --- | --- | --- | --- | --- | --- | --- | --- |
|  | OR | 95% CI | p | OR | 95% CI | p | OR | 95% CI | p | OR | 95% CI | p |
| Age | 1.01 | 0.98, 1.04 | 0.538 | 1.02 | 0.99, 1.05 | 0.182 | 1.03 | 0.99, 1.06 | 0.121 | 1.01 | 0.98, 1.04 | 0.538 |
| Gender (ref: male) |  |  |  |  |  |  |  |  |  |  |  |  |
| Female | 1.47 | 0.93, 2.33 | 0.098 | 1.65 | 1.03, 2.65 | 0.039 | 1.65 | 1.01, 2.70 | 0.047 | 1.47 | 0.93, 2.33 | 0.098 |
| Non-binary/other | 0.64 | 0.16, 2.57 | 0.529 | 0.98 | 0.24, 4.06 | 0.981 | 0.69 | 0.16, 3.02 | 0.621 | 0.64 | 0.16, 2.57 | 0.529 |
| Ethnicity (ref: White) |  |  |  |  |  |  |  |  |  |  |  |  |
| Asian/Asian British | 0.77 | 0.43, 1.38 | 0.381 | 0.59 | 0.32, 1.07 | 0.082 | 0.62 | 0.33, 1.16 | 0.134 | 0.77 | 0.43, 1.38 | 0.381 |
| Black/African/Caribbean/Black British | 1.74 | 0.71, 4.26 | 0.227 | 1.18 | 0.46, 3.03 | 0.731 | 1.40 | 0.54, 3.68 | 0.490 | 1.74 | 0.71, 4.26 | 0.227 |
| Mixed/multiple ethnic groups | 1.29 | 0.67, 2.48 | 0.455 | 1.24 | 0.61, 2.51 | 0.558 | 1.25 | 0.60, 2.57 | 0.552 | 1.29 | 0.67, 2.48 | 0.455 |
| Other | 0.42 | 0.14, 1.23 | 0.114 | 0.28 | 0.09, 0.86 | 0.026 | 0.29 | 0.09, 0.90 | 0.032 | 0.42 | 0.14, 1.23 | 0.114 |
| First language not English (ref: English) | 0.77 | 0.41, 1.42 | 0.401 | 0.66 | 0.35, 1.23 | 0.191 | 0.69 | 0.36, 1.32 | 0.262 | 0.77 | 0.41, 1.42 | 0.401 |
| International student (ref: home student) | 0.80 | 0.47, 1.37 | 0.419 | 0.97 | 0.55, 1.72 | 0.930 | 1.01 | 0.55, 1.86 | 0.968 | 0.80 | 0.47, 1.37 | 0.419 |
| Level of study (ref: undergraduate) |  |  |  |  |  |  |  |  |  |  |  |  |
| Masters | 1.16 | 0.71, 1.89 | 0.544 | 1.17 | 0.71, 1.93 | 0.536 | 1.06 | 0.64, 1.77 | 0.822 | 1.16 | 0.71, 1.89 | 0.544 |
| Doctoral | 1.68 | 0.59, 4.80 | 0.329 | 1.68 | 0.58, 4.87 | 0.344 | 1.62 | 0.55, 4.76 | 0.380 | 1.68 | 0.59, 4.80 | 0.329 |
| Academic field (ref: social science) |  |  |  |  |  |  |  |  |  |  |  |  |
| Humanities | **0.31** | **0.18, 0.55** | **<0.001** | 0.34 | 0.19, 0.62 | **<0.001** | 0.39 | 0.21, 0.72 | 0.003 | 0.31 | 0.18, 0.55 | **<0.001** |
| Natural sciences | 1.13 | 0.56, 2.29 | 0.727 | 1.12 | 0.55, 2.29 | 0.750 | 1.17 | 0.57, 2.41 | 0.675 | 1.13 | 0.56, 2.29 | 0.727 |
| Maths/computing (formal science) | **0.43** | **0.22, 0.82** | **0.010** | 0.42 | 0.22, 0.81 | 0.009 | 0.44 | 0.23, 0.87 | 0.018 | 0.43 | 0.22, 0.82 | 0.010 |
| Professions and applied sciences | 0.83 | 0.51, 1.35 | 0.445 | 0.83 | 0.50, 1.36 | 0.453 | 0.82 | 0.49, 1.36 | 0.442 | 0.83 | 0.51, 1.35 | 0.445 |
| Social needs | - | - | - | 1.02 | 0.93, 1.13 | 0.635 | 1.02 | 0.92, 1.13 | 0.700 | 1.02 | 0.92, 1.13 | 0.674 |
| Physical health problems or disabilities | - | - | - | **0.59** | **0.35, 1.00** | **0.051** | 0.64 | 0.37, 1.11 | 0.114 | 0.62 | 0.36, 1.08 | 0.094 |
| Mental health difficulties or conditions | - | - | - | **0.47** | **0.30, 0.72** | **0.001** | 0.36 | 0.21, 0.62 | **<0.001** | 0.36 | 0.21, 0.62 | **<0.001** |
| Mental wellbeing (SWEMWBS) | - | - | - | 1.03 | 0.98, 1.08 | 0.261 | 1.02 | 0.97, 1.07 | 0.456 | 1.02 | 0.97, 1.07 | 0.542 |
| Done things to look after mental health (ref: never) |  |  |  |  |  |  |  |  |  |  |  |  |
| Rarely | - | - | - | - | - | - | 0.70 | 0.23, 2.12 | 0.531 | 0.68 | 0.22, 2.09 | 0.506 |
| Sometimes | - | - | - | - | - | - | 1.29 | 0.44, 3.73 | 0.642 | 1.26 | 0.43, 3.70 | 0.668 |
| Often | - | - | - | - | - | - | 1.55 | 0.52, 4.64 | 0.435 | 1.53 | 0.51, 4.61 | 0.453 |
| Always | - | - | - | - | - | - | 0.98 | 0.29, 3.29 | 0.974 | 0.95 | 0.28, 3.21 | 0.934 |
| Spoken to professional about mental health | - | - | - | - | - | - | 0.78 | 0.47, 1.27 | 0.318 | 0.75 | 0.46, 1.23 | 0.257 |
| Other mental health support (ref: none) |  |  |  |  |  |  |  |  |  |  |  |  |
| Family/friend | - | - | - | - | - | - | 1.15 | 0.73, 1.83 | 0.551 | 1.15 | 0.73, 1.83 | 0.544 |
| Other | - | - | - | - | - | - | 1.08 | 0.30, 3.93 | 0.906 | 1.10 | 0.31, 3.94 | 0.888 |
| Taken medication for mental health | - | - | - | - | - | - | **1.93** | **1.09, 3.41** | **0.025** | 1.91 | 1.07, 3.38 | 0.027 |
| Engaged in social/community activities | - | - | - | - | - | - | 1.26 | 0.80, 1.96 | 0.314 | 1.21 | 0.77, 1.89 | 0.405 |
| Missed university due to mental health | - | - | - | - | - | - | 1.15 | 0.71, 1.88 | 0.565 | 1.15 | 0.71, 1.88 | 0.567 |
| Missed employed work due to mental health | - | - | - | - | - | - | 0.77 | 0.38, 1.56 | 0.466 | 0.72 | 0.35, 1.47 | 0.368 |
| Aware of SP previously | - | - | - | - | - | - | - | - |  | 1.43 | 0.86, 2.40 | 0.172 |

Note. N=692. OR: odds ratio. Bold text indicates p<0.05.

